# Bayesian Estimation of the Seroprevalence of Antibodies to SARS-CoV-2

**DOI:** 10.1101/2020.08.23.20180497

**Authors:** Qunfeng Dong, Xiang Gao

## Abstract

Accurately estimating the seroprevalence of antibodies to SARS-CoV-2 requires the use of appropriate methods. Bayesian statistics provides a natural framework for considering the variabilities of specificity and sensitivity of the antibody tests, as well as for incorporating prior knowledge of viral infection prevalence. We present a full Bayesian approach for this purpose, and we demonstrate the utility of our approach using a recently published large-scale dataset from the U.S. CDC.

## INTRODUCTION

Antibody tests for COVID-19 have been increasingly deployed to estimate the seroprevalence of antibodies to SARS-CoV-2^1^. Although antibody tests can provide important estimations on the prevalence of the viral infection in populations, the test results must be interpreted with caution due to the presence of false positives and false negatives^2^. Therefore, a critical statistical challenge is how to accurately estimate the prevalence of the viral infection in populations while accounting for the false positive and false negative rates of the antibody tests.

Recently, the U.S. Centers for Disease Control and Prevention (CDC) published a large-scale study on antibody tests from 10 sites in the U.S. administered between March 23 and May 12, 2020^3^. The CDC antibody tests employed an enzyme-linked immunosorbent assay with a specificity (i.e., 1 – false positive rate) of 99.3% (95% CI, 98.3%-99.9%) and sensitivity (i.e., true positive rate) of 96.0% (95% CI, 90.0%-98.9%)^3^. In order to take the test accuracy into the consideration, the CDC study applied the following simple correction: *R_obs_* = *P*×*Sensitivity* + (1-*P*) × (1-*Specificity*), where *R_obs_* is the observed seroprevalence in the study samples and *P* is the unknown seroprevalence in populations. Using the point estimates of the sensitivity (96.0%) and specificity (99.3%) of the antibody tests, they obtained the point estimate of the population prevalence *P* = (*R_obs_* – 0.007)/0.953.

There are two main limitations with such an approach. First, only the point estimate of population prevalence *P* was obtained. Although the CDC study also generated confidence intervals for the point estimate based on a non-parametric bootstrap procedure, the confidence interval does not provide a probabilistic measurement of the uncertainty associated with all possible values of the unknown prevalence. Second, the above CDC approach could not account for any prior knowledge of the population prevalence *P*, which can lead to inaccurate estimation especially when the true rate of viral infection is low, even with high specificity and sensitivity of the tests^4,5^.

To overcome the above limitations, we have developed a Bayesian approach. Our approach is not a simple application of Bayes’ theorem by plugging in the point estimates of sensitivity and specificity into the formula and computing a posterior probability. Instead, our approach is a full Bayesian procedure that models the known variability in the sensitivity (95% CI, 90.0%-98.9%) and specificity (95% CI, 98.3%-99.9%) of the antibody test, and we can incorporate any prior knowledge of the viral infection rate to estimate the entire posterior probability distribution of the unknown population prevalence.

## MATERIALS AND METHODS

### Bayesian modeling

Let *N_t_* and *N_p_* denote the number of people tested in total and the number of people tested as positive, respectively. Let *p* denote the unknown seroprevalence of antibodies to SARS-CoV-2. Let *θ* denote the true positive rate of the antibody test (i.e., sensitivity). Let *κ* denote the false positive rate of the test (i.e., 1 – specificity). Then, we can define the following likelihood function:

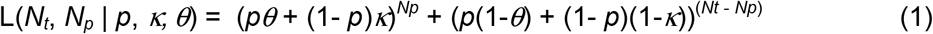

In Eq. (1), the term (*pθ* + (1-*p*)*κ*)*^Np^* corresponds to the probability of observing *N_p_* people that have tested positive, since a person with a positive test result can either be infected (with the probability of *p*) and correctly test positive (with the probability of *θ*), or not infected (with the probability of 1 -*p*) and falsely test positive (with the probability of *κ*). Similarly, the term (*p*(1-*θ*) + (1-*p*)*κ*)^(^*^Nt^*^-^*^Np^*^)^ corresponds to the probability of observing (*N_t_* -*N_p_*) people whose test results were negative.

To estimate the posterior probability of *p*,, we need to sample from the following posterior distribution:

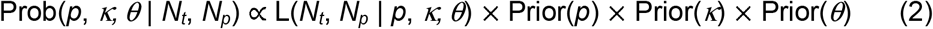

To specify the prior distribution for *p*, *κ*, and *θ*, we chose beta distributions as they are commonly used to model probabilities^6^.

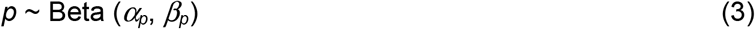

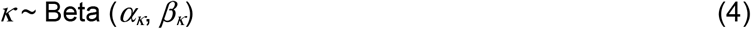

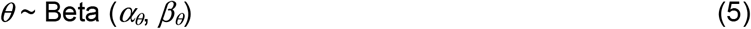

where *α_p_*, *β_p_*, *α_κ_*, *β_κ_*, *α_θ_*, and *β_θ_* denote shape parameters of the corresponding beta distributions. For the unknown parameter *p*, we chose to use a non-informative flat prior probability distribution for this study (i.e., *α_p_* = *β_p_* = 1), although it can be adjusted if prior knowledge of the proportion of infected people for a particular region is known (see more in the Discussion section). For *κ* and *θ*, we chose informative priors to reflect the known specificity and sensitivity of a particular antibody test. Specifically, the shape parameters of *α_κ_*, *β_κ_*, *α_θ_*, and *β_θ_* can be estimated using the method of moments^5^ as follows:

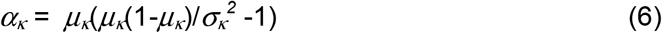

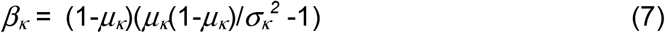

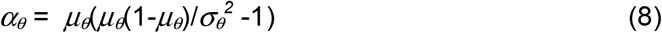

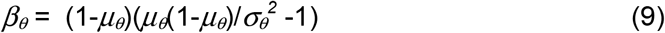

where *μ_κ_* and 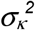, and *μ_θ_* and 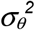 represent the mean and variance of the test specificity and sensitivity, respectively. For this study, the mean of specificity and sensitivity is 99.3% and 96.0%, respectively. The variances of specificity and sensitivity were approximated^7^ as *s(1-s)/n*, where *s* is the mean value of specificity or sensitivity, and *n* = 618 according to the CDC validation study on the antibody test accuracy^8^.

We used WinBUGS^9^ (version 1.4.3) to implement the above models. In particular, the likelihood function was implemented using the “*ones trick*”^10^ of WinBUGS (see the GitHub repository https://github.com/qunfengdong/AntibodyTest for the implementation details). The posterior distributions were estimated with the Markov Chain Monte Carlo (MCMC) sampling in WinBUGS using the following parameters: the number of chains of 4, the number of total iterations of 100,000, burn-in of 10,000, and thinning of 4. Convergence and autocorrelations were evaluated with trace/history/autocorrelation plots and the Gelman-Rubin diagnostic^11^. Multiple initial values were applied for MCMC sampling. The above Bayesian procedure was validated with simulated datasets generated by our customized R^12^ script (available in the above GitHub repository).

### Seroprevalence data

The seroprevalence data was taken from the aforementioned CDC publication^3^. Our approach requires two inputs: (i) the total number of tested samples and (ii) the number of positive samples. For this project, we only focused on gender-specific data in the CDC study. We extracted the total number of male and female samples from the original Table 1 in the CDC publication. However, the number of positive samples was not reported in the CDC publication. To infer those numbers for both genders, we extracted the CDC estimated seroprevalence, *P*, for both genders from the original Table 2 in the CDC publication. Using the equation *P* = (*R_obs_* – 0.007)/0.953 mentioned above, we obtained the observed seroprevalence *R_obs_* for both genders, which were used for calculating the number of observed positive male and female samples by multiplying *R_obs_* to the total number of samples in each respective gender. Table 1 lists the calculated number of test positive samples, rounded to the nearest integer in each site.

**Table 1.**
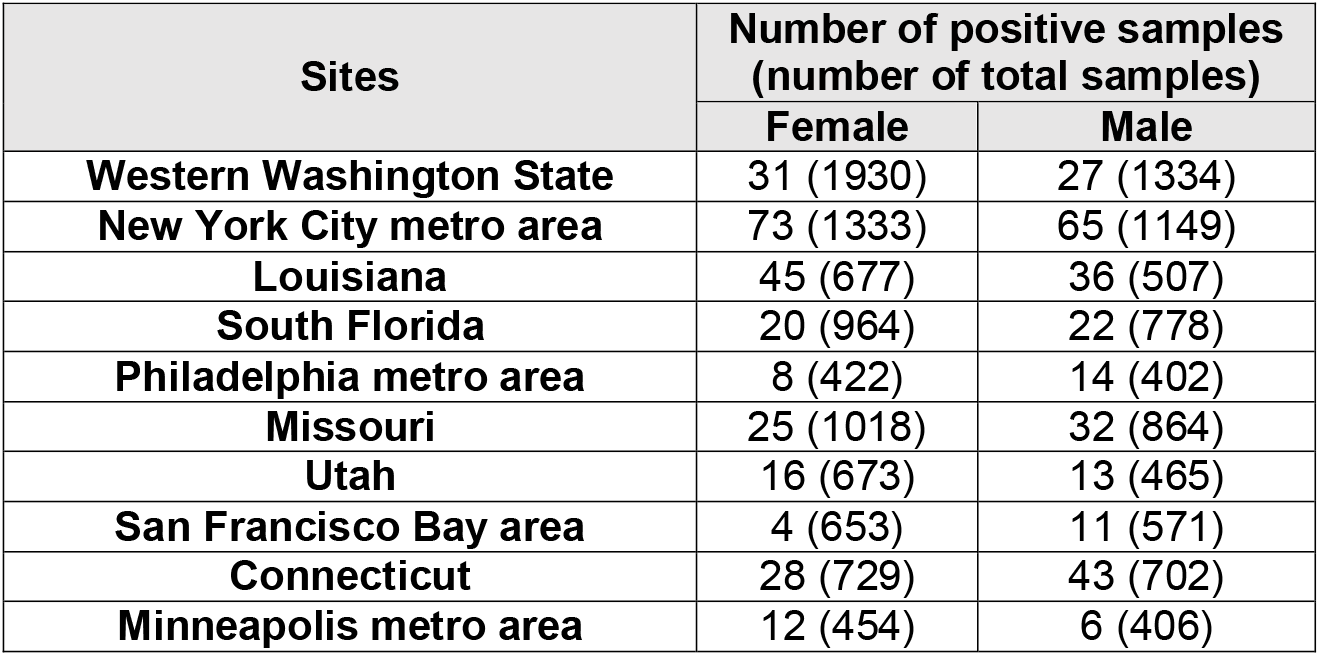
Number of positive samples calculated from the CDC publication^3^.

## RESULTS

We applied our Bayesian approach to the data listed in Table 1. It is important to emphasize that Bayesian approaches produce entire probability distributions instead point estimates^6^. Figure 1 depicts the posterior distributions of the seroprevalence of antibodies to SARS-CoV-2 virus in 10 U.S. sites. Table 2 lists both the original CDC point estimates with the accompanying 95% confidence intervals, and our Bayesian estimates, which were presented as the medians and 95% credible intervals of the posterior distributions. It is worth noting that confidence intervals and Bayesian credible intervals are two different concepts^13^, thus they are not technically comparable despite being listed together in Table 2 for convenience. Although the posterior medians are similar to the original CDC point estimates overall, the entire posterior distributions (fig. 1) inferred by our Bayesian approach accurately capture the uncertainties associated with seroprevalence (i.e., the posterior distribution provides a precise probability associated with every possible value of seroprevalence), which cannot be achieved through confidence intervals.

**Figure 1.**
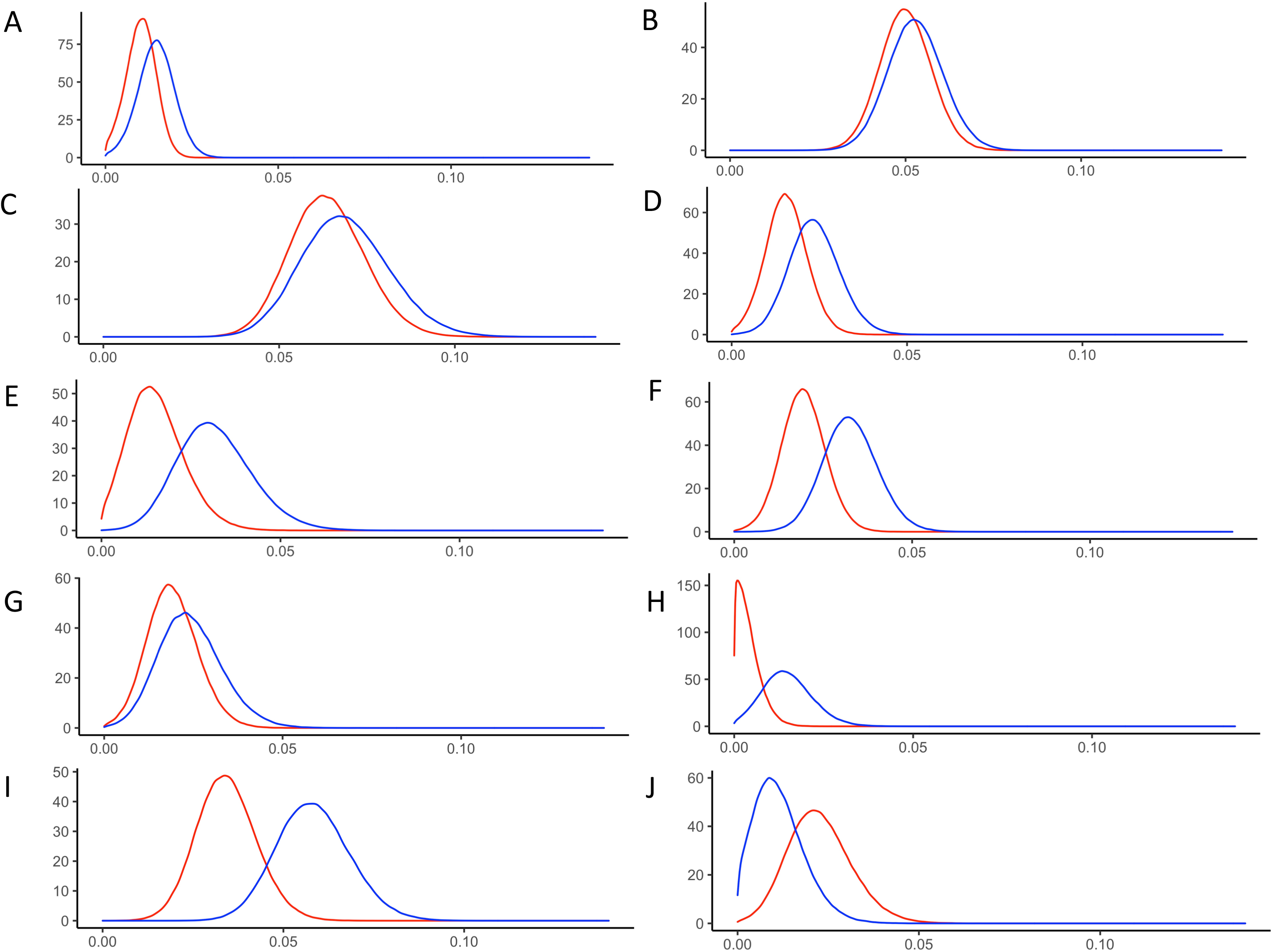
The posterior probability density of the prevalence of female (red) and male (blue) infected by SARS-CoV-2 virus in 10 U.S. sites: **(A)** Western Washington State, **(B)** New York City metro area, **(C)** Louisiana, **(D)** South Florida, **(E)** Philadelphia metro area, **(F)** Missouri, **(G)** Utah, **(H)** San Francisco Bay area, **(I)** Connecticut, and **(J)** Minneapolis metro area.

**Table 2.** Estimated seroprevalence of antibodies to SARS-CoV-2 in populations.

| Sites | CDC estimate<sup>3</sup><br>(95% confidence interval), % |  | Posterior median<br>(95% credible interval), % |  |
| --- | --- | --- | --- | --- |
|  | Female | Male | Female | Male |
| **Western Washington State** | 1.7<br>(0.7-1.9) | 1.4<br>(0.8-2.4) | 1.0<br>(0.2-1.9) | 1.5<br>(0.4-2.5) |
| **New York City metro area** | 5.7<br>(4.2-7.0) | 5.9<br>(4.5-7.6) | 5.0<br>(3.6-6.5) | 5.3<br>(3.8-6.9) |
| **Louisiana** | 7.0<br>(4.7-9.4) | 6.8<br>(4.2-9.3) | 6.3<br>(4.4-8.6) | 6.8<br>(4.6-9.5) |
| **South Florida** | 2.2<br>(1.2-3.4) | 2.2<br>(1.1-3.6) | 1.5<br>(0.4-2.8) | 2.3<br>(1.0-3.8) |
| **Philadelphia metro area** | 1.9<br>(0.7-3.7) | 3.0<br>(1.3-5.2) | 1.5<br>(0.2-3.2) | 3.1<br>(1.3-5.4) |
| **Missouri** | 2.6<br>(1.5-3.7) | 3.1<br>(1.8-4.6) | 1.9<br>(0.7-3.2) | 3.2<br>(1.8-4.8) |
| **Utah** | 2.5<br>(1.2-4.1) | 2.2<br>(0.9-3.6) | 1.9<br>(0.6-3.4) | 2.4<br>(0.8-4.3) |
| **San Francisco Bay area** | 0.7<br>(0.2-1.9) | 1.2<br>(0.4-2.7) | 0.3<br>(0.02-1.2) | 1.4<br>(0.3-3.0) |
| **Connecticut** | 4.1<br>(2.6-5.9) | 5.7<br>(3.8-7.6) | 3.4<br>(1.9-5.1) | 5.8<br>(3.9-7.9) |
| **Minneapolis metro area** | 2.7<br>(1.2-4.8) | 0.7<br>(0-2.3) | 2.2<br>(0.7-4.2) | 1.1<br>(0.1-2.7) |

## DISCUSSION

Antibody tests have been increasingly applied to estimate the prevalence of people who have been infected by the SARS-CoV-2 virus. For example, New York City recently released data of more than 1.46 million coronavirus antibody test results on August 18, 2020. Accurately analyzing such data is critical for developing important public health policies^14^. Our Bayesian approach can account for the variabilities in antibody tests (i.e., uncertainties in the sensitivity and specificity of the tests). In addition, the Bayesian approach can easily incorporate prior knowledge of the proportion of infected people for a particular region. This is particularly important for accurate estimation if the true prevalence is low^5^. Moreover, the Bayesian approach also provides a natural framework for updating the estimation based on new data, which is particularly relevant to the continuous monitoring of the seroprevalence of coronavirus antibodies. For example, New York City is still releasing coronavirus antibody test results on a weekly basis^15^. By turning the estimated posterior distribution from previous weeks into a prior distribution for the next week, the seroprevalence of coronavirus antibody can be quickly updated within a solid Bayesian probabilistic inference framework.

## Data Availability

We only used published dataset for analysis in this study.

## AUTHOR CONTRIBUTION

Q.D. and X.G. both contributed project conception. Q.D. contributed WinBUGS modeling and drafting the manuscript. X.G. contributed R programming and data analysis.

